# Persisting pulmonary dysfunction in pediatric post-acute Covid-19

**DOI:** 10.1101/2022.02.21.22270909

**Authors:** Rafael Heiss, Alexandra Wagner, Lina Tan, Sandy Schmidt, Adrian P. Regensburger, Franziska Ewert, Dilbar Mammadova, Adrian Buehler, Jens Vogel-Claussen, Andreas Voskrebenzev, Manfred Rauh, Oliver Rompel, Armin M. Nagel, Simon Lévy, Sebastian Bickelhaupt, Matthias S. May, Michael Uder, Markus Metzler, Regina Trollmann, Joachim Woelfle, Ferdinand Knieling

**Affiliations:** Institute of Radiology, University Hospital Erlangen, Friedrich-Alexander-Universität (FAU) Erlangen-Nürnberg, Germany; Department of Pediatrics and Adolescent Medicine, University Hospital Erlangen, Friedrich-Alexander-Universität (FAU) Erlangen-Nürnberg, Germany; Center for Social Pediatrics, University Hospital Erlangen, Friedrich-Alexander-Universität (FAU) Erlangen-Nürnberg, Germany; Translational Experimental and Molecular Imaging Laboratory (PETI_Lab), Department of Pediatrics and Adolescent Medicine, University Hospital Erlangen, Friedrich-Alexander-Universität (FAU) Erlangen-Nürnberg, Germany; Institute for Diagnostic and Interventional Radiology, Hannover Medical School, Hannover, Germany

**Author notes:** corresponding author Correspondence: PD Dr. med. habil. Ferdinand Knieling, Departments of Pediatrics and Adolescent Medicine, University Hospital Erlangen, Friedrich-Alexander-Universität (FAU) Erlangen-Nürnberg. shared authorship (Rafael Heiss, Alexandra Wagner, Lina Tan).

## Abstract

The frequency and extent of persistent sequelae in children and adolescents after infection with SARS-CoV-2 still needs to be comprehensively determined. In this cross-sectional clinical trial, we used non-invasive, label-free morphologic and free-breathing phase-resolved functional low-field magnetic resonance imaging (LF-MRI) to identify pulmonary changes in children and adolescents from 5 to <18 years after previously PCR-confirmed SARS-CoV-2 infection. While morphological pathologies were less frequent in children, functional LF-MRI visualized widespread ventilation, perfusion and combined ventilation/perfusion defects compared to healthy controls. The loss of functional lung parenchyma was more pronounced in long Covid than recovered patients. While pulmonary dysfunction was persistent even month after primary infection, LF-MRI demonstrated high capability to visualize and detect these changes in children and adolescents. (Clinicaltrials.org ID NCT04990531)

## Introduction

SARS-CoV-2 has emerged as a global pandemic causing more than 280 million documented infections and 5.4 million deaths until the end of 2021.^1^ In comparison to adults, there is common evidence that Covid-19 in children and adolescents has a milder course with recovery within weeks.^2^ This finding is compromised by a growing body of evidence for post-acute sequelae and symptoms in these age classes.^2-4^ While there is an increasing understanding of the multi-organ damage of Covid-19 beyond the acute phase of infection^5^, the nature, frequency and definition of post-acute sequelae in children and adolescents still remains undetermined with a discrepancy in clinical appearance and objective findings.^6^ A major proportion of pediatric studies have lately prioritized research in mental health issues during the Covid-19 pandemic^7-10^, while other studies have already raised concerns on ongoing disease manifestations, including increased thrombotic state, microangiopathy and inflammation.^11-13^

As the lung is a primary target of the SARS-CoV-2 virus,^14^ computed tomography aided in the diagnosis of pulmonary manifestation of Covid-19 in adults.^15^ Even three months after infection, angiographic imaging of pulmonary microcirculation still revealed widespread microangiopathy in over 65% of the patients.^16^ Such techniques using invasive procedures or ionization radiation are not feasible in children and also seem to have limited diagnostic value as lung parenchymal changes do present less obvious and pronounced.^17-19^ Therefore, there is an unmet clinical need to more precisely characterize pulmonary manifestations in children and adolescents after SARS-CoV 2 infection.

We used low-field magnetic resonance imaging (LF-MRI) for imaging of the pediatric lung. At low field strength, this technique has improved imaging quality of near air-tissue interfaces, without the need for ionizing radiation.^20,21^ The aim of the study was to characterize both morphologic and functional changes of lung parenchyma in PCR-proven SARS-CoV-2 pediatric post-acute Covid-19 patients in comparison to healthy controls.

## Methods

### Trial design

We performed a single-center, cross-sectional, investigator-initiated trial to investigate lung parenchymal changes in children and adolescents after SARS-CoV-2 infection. The call for study participation for Covid-19 patients was nationwide public and consecutive. After assessment for clinical parameters, a blood sample was drawn and all patients underwent LF-MRI. Clinical features during and after infection, the time period from positive PCR test and laboratory parameters were compared to imaging results. Details are provided in the protocol and the statistical analysis plan.

The coordinating clinical investigators were responsible for data collection and site monitoring. The first authors and corresponding author had constant access to the data and performed the statistical analysis as well as the creation of the first draft of the manuscript independently from any commercial facility.

### Participants

All participants in both cohorts were between 5 and <18 years of age. In the Covid-19 group, eligible patients required a positive PCR-test for SARS-CoV-2 in the prior history. The definition for long Covid was based on the persistence of symptoms for a minimum of 12 weeks and either one of the four criteria:^22,23^ 1) Symptoms that persist from the acute Covid-19 phase or its treatment, 2) Symptoms that have resulted in a new health limitation, 3) New symptoms that occurred after the end of the acute phase but are understood to be a consequence of Covid-19 disease, 4) Worsening of a pre-existing underlying condition. In the healthy control group, children required a negative medical history, post-hoc negative serological antibody status, and no prior immunization against SARS-CoV-2.

### Outcome measures

The primary outcome was the determination of the frequency of morphological changes of lung parenchyma by LF-MRI. Secondary outcomes included functional lung changes comprising ventilation defects (ventilation defected percentage; VDP_Total_), perfusion defects (perfusion defected percentage; QDP_Total_), the match (ventilation/perfusion match; VQM_Non-defect_) and match (defect) of both (ventilation/perfusion defect; VQM_Defect_), laboratory assessments and anamnestic clinical symptoms.

### Procedures

#### Clinical data and blood samples

Patients were assessed for medical history, including symptoms during and after Covid-19 infection. Blood pressure and heart rate were measured from each individual. A blood sample was collected to assess blood count, interleukin 6 (IL-6), C-reactive protein (CrP) and antibodies against SARS-CoV-2 (spike protein and nucleocapsid antibodies, electrochemiluminescence immunoassay (ECLIA) Elecsys® Anti-SARS-Cov-2 S and Anti-SARS-CoV2, Roche, Switzerland); for details see Methods section in Supplementary Appendix).^24^

#### Magnetic resonance imaging

##### Imaging setup

All participants underwent morphological and functional low-field magnetic resonance imaging (LF-MRI, 0,55 Tesla MAGNETOM Free.Max, Siemens Healthineers, Erlangen, Germany) for visualization of morphological features, ventilation and perfusion of the lung.^25-29^ For all investigations a standard body coil was used for free-breathing lung imaging. The final parameters were: one two-dimensional central coronal slice positioned at the middle of the lung hila, thickness=15mm, in-plane resolution=1.7×1.7mm^2^, matrix=128×128 (interpolated to 256×256), bandwidth=1149Hz/pixel, flip angle=80°, TR/TE=292.8/1.6ms, parallel imaging acceleration factor =2, no partial Fourier, 250 time points, temporal resolution=300ms, duration=1min15s. The evaluating radiologist was blinded to clinical features for all analyses.

##### Morphologic lung imaging

For morphological lung assessment, a coronal and transversal Turbo-Spin-Echo (TSE) sequence with BLADE (periodically rotated overlapping parallel lines with enhanced reconstruction) readout and respiration-gating were acquired. The coronal image was acquired with a STIR preparation (Short-Inversion Time Inversion Recovery), T2-weighting (TE/TR=74/2500ms), 1.5×1.5mm^2^ in-plane resolution, 272×272 matrix, 6mm thickness. The transversal image was proton density-weighted (TE/TR=33/2000ms) with 1.3×1.3mm^2^ in-plane resolution, 304×304 matrix, 6mm thickness.

##### Functional lung assessment

For free-breathing phase-resolved functional lung (PREFUL) LF-MRI, the following parameters were calculated voxel-wise by using a dedicated software (MR Lung v2.0, Siemens Healthcare, Erlangen, Germany) after automatic registration to a mid-expiration position and lung parenchyma segmentation. ^26^

Normalized perfusion (Q, %) with respect to a full-blood signal region, determined as the highest perfusion signal region in-between the lungs and expected to reflect the aorta or other available large vessel ^27^; Regional ventilation (V, %) calculated as: 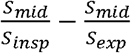 with S the signal value at end-inspiration (*insp*), end-expiration (*exp*) and middle position (*mid)*; ^30^ Flow-Volume Loop correlation (FVL): correlation of the Flow-Volume Loop (deduced from the V reconstructed cycle) with respect to a healthy region (largest connected region within the 80^th^ and 90^th^ V percentiles).^31^ Based on those maps, the percentage of defect areas (QDP_Total_, VDP_Total_) were calculated based on thresholds optimized on a large cohort (of 155 healthy volunteers and 95 patients with various lung diseases scanned at 1.5T with a Fast Low-Angle Shot sequence (perfusion: 2%, FV: 40% of the 90th percentile, FVL: 0.9) (not published yet). The percentage of concurrent defect areas of perfusion and ventilation metrics (VQM_Defect,_ VQM_Defect, FVL_) and perfusion defects exclusive to ventilation defects (QDP_Exclusive_) were derived, and vice versa (VDP_FVL,Exclusive_, VDP_Exclusive_). In addition, areas without defects on both perfusion and ventilation maps (VQM_Non-defect_, VQM_Non-defect, FVL_) were calculated. An overview and explanation of all parameters used is given in **Table S1**. PREFUL MRI ventilation and perfusion measures were recently validated using V/Q single photon emission tomography, dynamic contrast enhanced MRI as well as ^19^F and ^129^Xe inhaled gas MRI.^27,32^

### Statistical analyses

Continuous variables are given as mean value with standard deviation, categorical variables as numbers with percentages. The occurrence of MRI changes is given as a percentage of the population. A non-parametric Mann-Whitney test was used for pairwise comparisons. A non-parametric Kruskal-Wallis test with Dunn’s test for correction of multiple testing was used to assess differences in healthy controls, recovered and long Covid patients. Receiver operator characteristic (ROC) curves were calculated using healthy controls (SARS-CoV-2 PCR and antibody negative) vs. recovered patients (SARS-CoV-2 PCR positive; no persisting symptoms) or long Covid patients (SARS-CoV-2 PCR positive; persisting symptoms). Prism 9, version 9.3.1 (GraphPad Software, California, USA) was used for all statistical analyses. A P value of less than 0.05 was considered to indicate statistical significance in all analyses.

## Results

### Patient population

Between August 9, 2021 and December 30, 2021, a total of 17 healthy controls and 91 pediatric patients after PCR-positive SARS-CoV-2 infection were screened. 7 healthy controls and 33 post-acute Covid-19 patients were excluded prior to participation. 1 healthy control had a pulmonary nodule of unknown dignity on morphologic MRI and was therefore excluded. 4 post-acute Covid-19 patients were not able to complete LF-MRI scanning and were therefore excluded. Overall, 54 post-acute Covid-19 patients and 9 controls completed clinical, laboratory and LF-MRI assessments (**Figure 1**).

**Figure 1.**
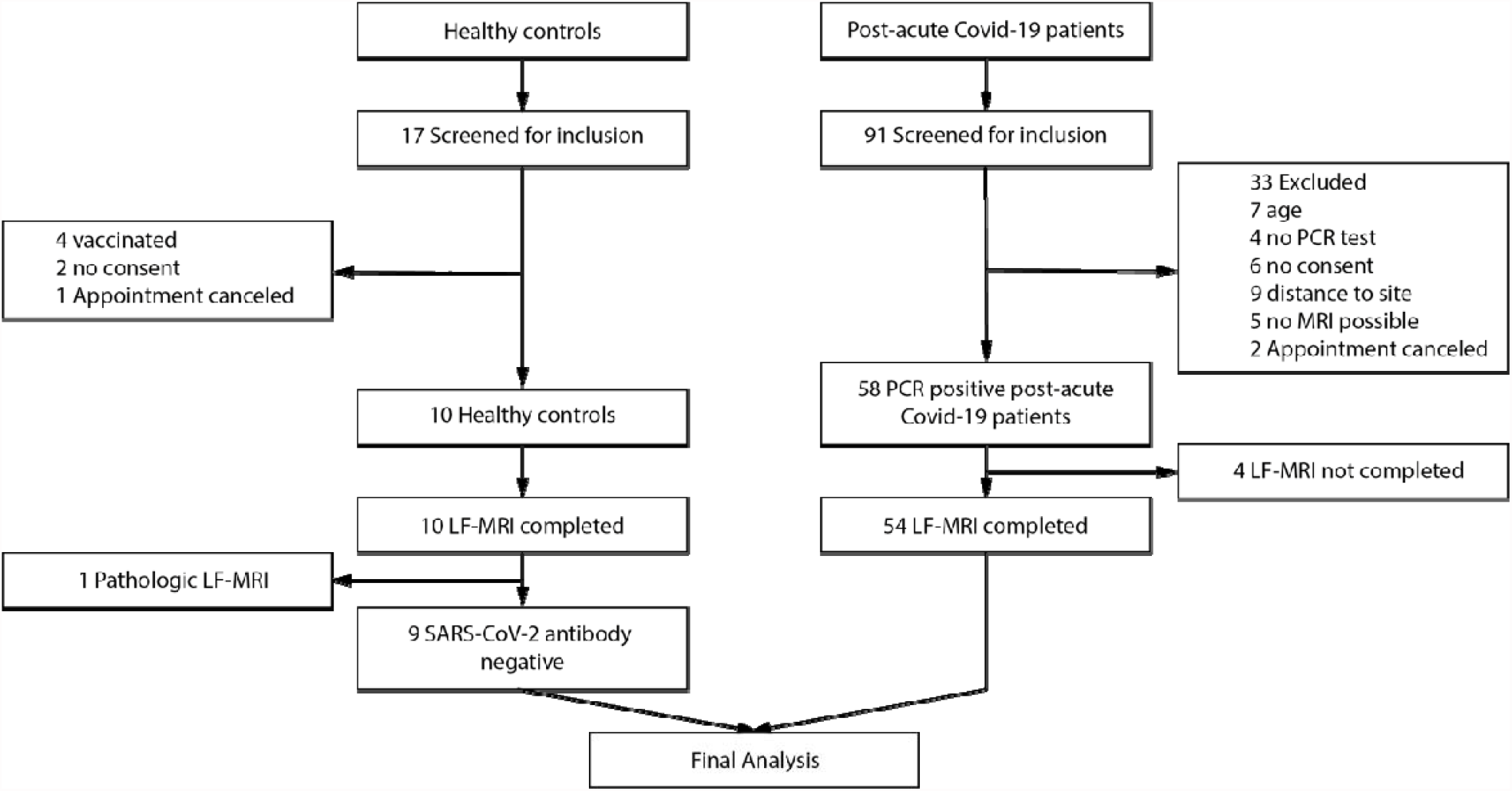
Flow chart of the study.

The characteristics of the participants were similar in both groups (**Table 1**). The mean age of post-acute Covid-19 patients was 11.5±3.2 years (control: 10.3±3.1 years), mean weight was 47.6±17.9kg (39.7±15.3kg), mean height was 155.7±17.3 (143.9±11.3cm) and 44% (30%) were female. 25 patients (46% of all patients) were classified as having long Covid. 5 patients reported headache (9%), 15 dyspnea (28%), 1 pneumonia (2%), 4 anosmia (7%), 1 ageusia (2%), 4 fatigue (7%), 6 impaired attention (11%), 1 limb pain (2%) and 16 shortness of breath (30%) (**Table S1**). Detailed acute and post-acute Covid-19 symptoms can be found in **Table S2**. 4 patients with PCR-positive SARS-CoV2 infection did not show any symptoms during acute infection. None of the Covid-19 patient group required hospital admission during the primary infection period. The median interval between positive SARS-CoV-2 PCR test and study participation was 222±134 days. There were no missing primary and/or secondary outcome data.

**Table 1.** Demographic and Clinical Characteristics of the Participants*.

| **Characteristic** | **Healthy control**<br>**(N=9)** | **Post Covid-19**<br>**(N=54)** |
| --- | --- | --- |
| **Age – yr†** | 10.3±3.1 | 11.5±3.2 |
| **Age group – no. (%)** |  |  |
| 5-8 yr | 3 (33%) | 10 (19%) |
| 8-12 yr | 4 (44%) | 24 (44%) |
| 12-18 yr | 2 (22%) | 20 (37%) |
| **Weight – kg** | 39.7±15.3 | 47.6±17.9 |
| **Height – cm** | 143.9±11.3 | 155.7±17.3 |
| **Sex – no. (%)** |  |  |
| Female | 3 (30%) | 24 (44%) |
| Male | 7 (70%) | 30 (56%) |
| **Elapsed time from positive PCR – days** | n/a | 222±134 |
| **Vaccination prior to infection – no. (%)** | n/a | 1 (2%) |
| **Vaccination prior to inclusion – no. (%)** | 0 (0%) | 16 (30%) |
| **Hospitalization after SARS-CoV-2 infection** | n/a | 0 (0%) |
| **Long Covid symptoms – no. (%)** | n/a | 25 (46%) |
| **Vital parameters** |  |  |
| Systolic blood pressure - mmHg | 114±12 | 115±13 |
| Diastolic blood pressure - mmHg | 64±5 | 68±9 |
| Heart rate – bpm | 84±11 | 85±15 |
| **Laboratory assessments** |  |  |
| Hemoglobin – g/dl | 13.3±0.8 | 13.5±1.2 |
| Thrombocytes – *10<sup>3</sup>/μl | 309±61 | 287±62 |
| Leukocytes – *10<sup>3</sup>/μl | 6.6±1.5 | 6.7±1.6 |
| Interleukin 6 – ng/l | 0.7±1.5 | 0.2±0.8 |
| C-reactive protein – mg/l | 0.5±0.5 | 0.5±0.9 |
| **SARS-CoV-2 serostatus** |  |  |
| Spike protein antibody — U/ml‡¶ | 0±0 | 5846±13833 |
| Seropositive patients – No. (%) | 0 (0) | 50 (93) |
| Nucleocapsid antibody — U/ml‡¶ | 0±0 | 56±71 |
| Seropositive patients – No. (%) | 0 (0) | 48 (89) |

\* Plus-minus values are means ±SD.
† Age reflects the age at the date of signed informed consent.
‡ Serostatus at baseline was measured by Elecsys® Anti-SARS-Cov-2 S and Anti-SARS-CoV2 (Roche, Switzerland).

### Primary and secondary outcomes

Of the 54 post-acute Covid-19 and 9 healthy controls scanned with LF-MRI, 1 recovered Covid-19 patient showed morphological changes with pulmonary consolidations (**Figure S1**). All other patients did not show morphologic changes such as signs of pneumonia, infiltrations, ground-glass opacifications or fibrosis.

When compared to healthy controls, an increase in ventilation defects (VDP_Total_; 12.8±3.6% vs. 23.3±9.0, P<0.001), perfusion defects (QDP_Total_; 6.5±5.0% vs. 20.5±18.7%, P=0.05) and combined defects (VQM_Defect_; 0.5±0.8 vs. 4.6±5.7, P=0.001) was found in diseased patients using functional LF-MRI. Unaffected lung parenchyma was reduced from 81.2±6.1% in healthy volunteers to 60.8±19.1% (P<0.001) in post Covid patients (**Figure S2**).

When separating Covid-19 patients by clinical characteristics, VDP_Total_ increased from 12.8±3.6% in healthy controls to 22.1±8.1% (P=0.01) in recovered patients to 24.6±10.0% (P=0.002) in long Covid patients. Similarly, QDP_Total_ was increased from 6.5±5.0% in healthy controls to 19.5±19.1% (P=0.35) in recovered to 21.6±18.6% (P=0.10) in long Covid patients. Combined ventilation/perfusion defects (VQM_Defect_) increased from 0.5±0.8% to 3.9±4.7% (P=0.04) to 5.4±6.6% (P=0.002). Normal functional lung parenchyma was reduced from 81.2±6.1% in healthy controls to 62.0±18.7% (P=0.006) in recovered to 59.9±19.8 (P=0.003) in long Covid patients (**Table 2, Table S3** in the Supplementary Appendix for further parameters). Representative functional LF-MRI images are shown in **Figure 2;** the corresponding morphologic images are shown in **Figure S3**. For a complete imagimg list displaying combined ventilation/perfusion defects see **Figures S4-11** in the Supplementary Appendix.

**Table 2.** Quantitative measurements of free breathing phase-resolved functional lung magnetic resonance imaging*.

| **Parameter** | **Healthy control**<br>**(N=9)** | **Recovered**<br>**(N=29)** | **P**<br>**Value†** | **Long Covid**<br>**(N=25)** | **P**<br>**Value†** |
| --- | --- | --- | --- | --- | --- |
| Mean Perfusion - % | 6.5±1.6 | 5.2±2.3 | 0.22 | 5.0±2.3 | 0.07 |
| Mean Ventilation - % | 13.7±3.4 | 14.2±3.9 | >0.99 | 15.5±6.5 | >0.99 |
| Ventilation defected<br>percentage (VDP<sub>Total</sub>) -<br>% | 12.8±3.6 | 22.1±8.1 | 0.01 | 24.6±10.0 | 0.002 |
| Perfusion defected<br>percentage (QDP<sub>Total</sub>) -<br>% | 6.5±5.0 | 19.5±19.1 | 0.35 | 21.6±18.6 | 0.10 |
| Ventilation/perfusion<br>match defect (VQM<sub>Defect</sub>)<br>- % | 0.5±0.8 | 3.9±4.7 | 0.04 | 5.4±6.6 | 0.002 |
| Ventilation/perfusion<br>match<br>(VQM<sub>Non-defect</sub>) - % | 81.2±6.1 | 62.0±18.7 | 0.006 | 59.9±19.8 | 0.003 |

\* Plus-minus values are means ±SD.
† P Values tested with Kruskal-Wallis test and Dunn's post test for multiple comparisons and given in comparison to healthy controls.

**Figure 2.**
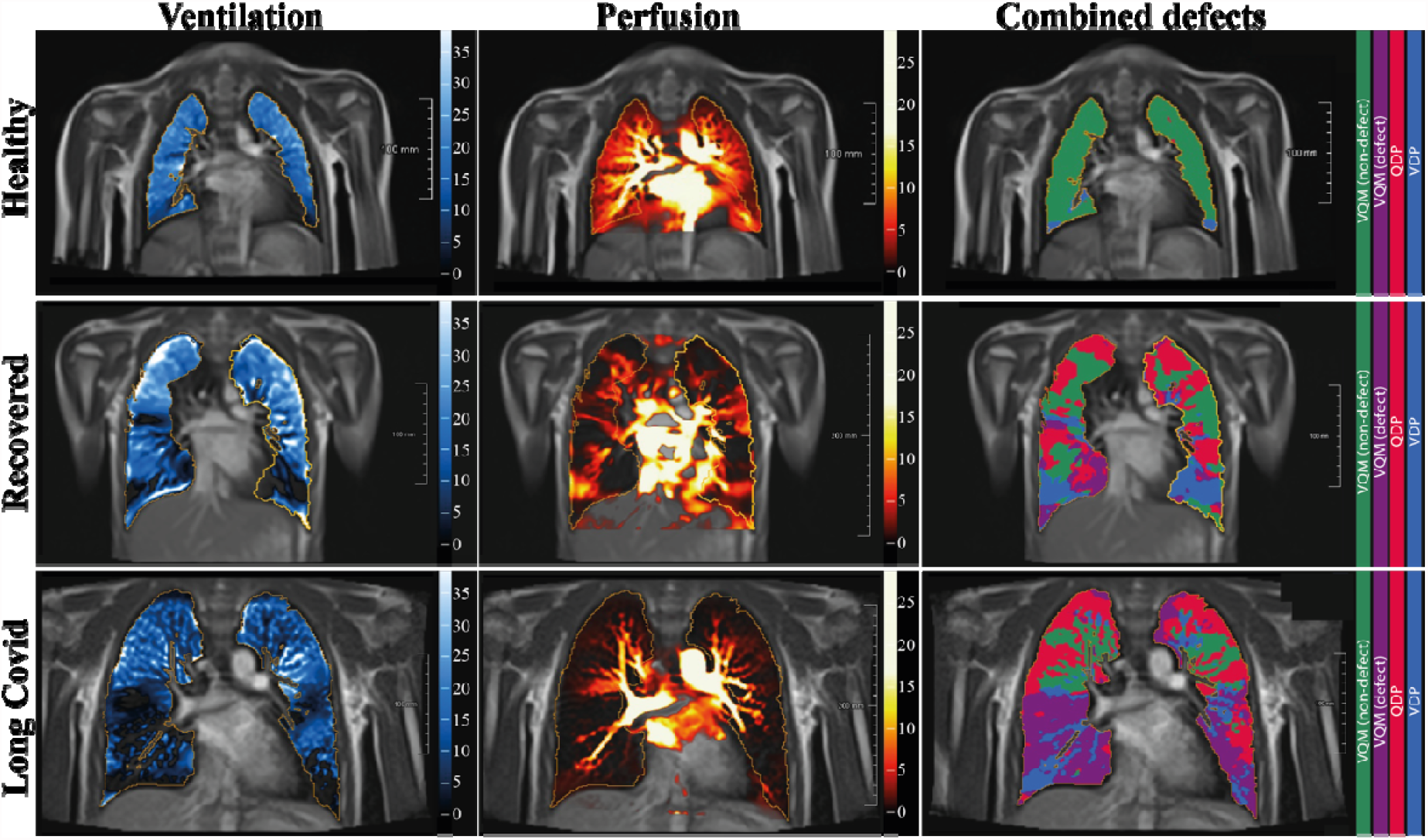
Functional low-field magnetic resonance imaging of pediatric post-acute Covid-19. From left to right: Representative functional low-field magnetic resonance images for ventilation defects, perfusion defects and combined defects in a healthy control (upper row), a recovered patient (middle row) and a long Covid patient (lower row).

A receiver operating characteristics (ROC) curve, performed for VDP_Total_, QDP_Total_, VQM_Defect_ and VQM_Non-defect_ using PCR proven infection as reference is shown in **Figure 3**. Post-acute Covid-19 patients could be differentiated from healthy controls with more pronounced differences in long Covid compared to recovered patients.

**Figure 3.**
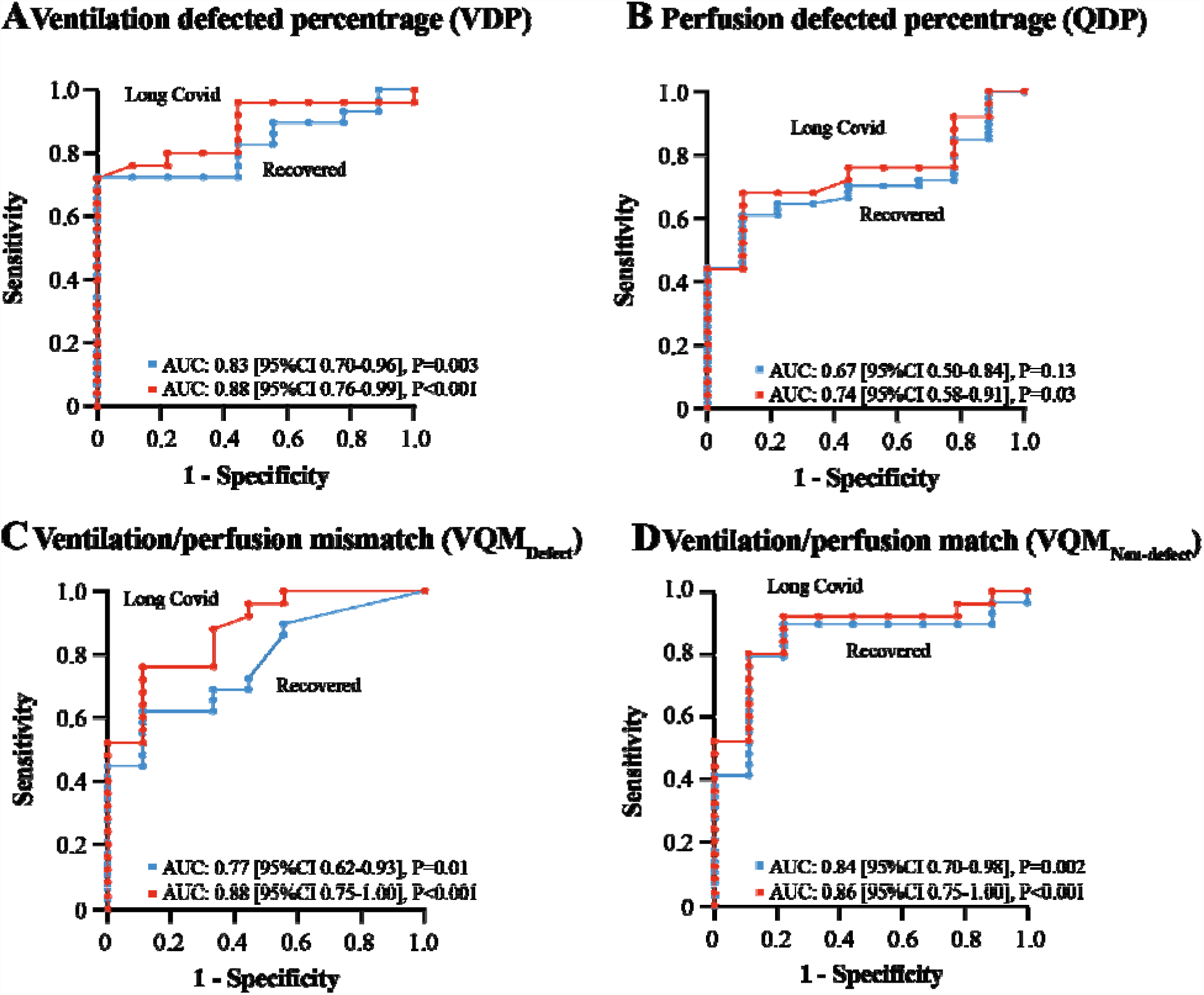
Diagnostic performance of low-field magnetic resonance imaging in pediatric healthy controls (N=9), recovered patients (N=29) and long Covid patients (N=25) Receiver operator characteristics (ROC) curve for ventilation defects, perfusion defects, ventilation/perfusion match (non-defect) and ventilation/perfusion match (defect) in recovered (n=29) and long Covid (n=25) patients versus healthy controls (n=9).

For VDP_Total_, the AUC was 0.83 [95%CI: 0.70-0.96] (P=0.003) in recovered and 0.88 [95%CI: 0.76-0.99] in long Covid patients. For QDP_Total_ the differences were less pronounced (0.67 and 0.74), while the VQM_Defect_ demonstrated a similar trend (0.77 and 0.88). Best differentiation was seen for VQM_Non-Defect_ with an AUC of 0.84 [95%CI: 0.70-0.98] (P=0.002) for recovered and 0.88 [95%CI: 0.75-1.00] (P<0.001) for long Covid patients. Using a cut-off of <77.4% 47 of 54 (87%) of the patients were classified as having a pathologic decrease in functional lung parenchyma on LF-MRI (**Table S4, Figure S12**).

### Laboratory assessments

From the 54 post-acute Covid-19 patients, 4 showed negative spike protein as well as nucleocapsid antibody level at time of presentation (time range to PCR-proven infection 186-416 days). 2 post-acute Covid-19 patients had reactive spike protein antibodies without reactive nucleocapsid antibodies (time range to PCR-proven infection 40-339 days). All 9 healthy controls were confirmed with negative spike protein and nucleocapsid antibody levels.

Inflammation parameters including CrP, IL-6 and blood counts were not suggestive of a current infection at the day of study for any participant.

## Discussion

In this monocentric diagnostic trial, we used LF-MRI for morphologic and functional characterization of lung parenchyma after pediatric SARS-CoV-2 infection. The trial included a heterogeneous population of children and adolescents with regard to acute or post-acute symptoms and time interval since primary infection. When compared to healthy controls, both, recovered and long Covid patients showed an increase in ventilation and perfusion defected lung parenchyma.

Similar imaging approaches to the one used in our study have already proven to be able to visualize pathologic changes in healthy pulmonary hypertension, cystic fibrosis or chronic obstructive pulmonary disease.^27-29^ Specifically for Covid-19, our findings tie up closely with observations in adults, where vascular^16^ or structural abnormalities^33^ persist in previously hospitalized adult patients. Another approach to assess lung function using inhaled gas contrasts agents that has already been successfully applied in adult Covid-19 patients, is hyperpolarized ^129^Xe MRI. Herein damaged lung gas exchange function^34^ and capillary diffusion limitation^35^ have been demonstrated. While using contrast agents might be less feasible in pediatric patients, results of ^129^Xe MRI in adults tie up with the findings of this first LF-MRI study in children and adolescents. In contrast to the consensus to date, assuming less severe Covid-19 infections and sequelae in younger patients, our study demonstrates widespread functional lung alterations are present in children and adolescents. This expands the understanding of pediatric post-acute Covid-19 disease, with the relevance of our findings even increasing as SARS-CoV-2 incidence is rising in most countries.^36^

Pathophysiology of acute and post-acute pathology of Covid-19 partly originates from direct endothelial damage, local inflammation and prothrombotic milieu.^14,37-39^ A proposed mechanism is the ACE2-mediated entry of SARS-CoV-2, which allows the virus to directly invade endothelial cells.^14,40^ This may explain manifestations such as pulmonary microangiopathy and widespread capillary microthrombi seen in autopsies from patients who died from Covid-19^14^ and fibrotic-like consolidations found in computed tomography.^33^ Previously described^13^ persisting signs of inflammatory processes could not be confirmed in our study. Putting into context, that children develop a robust, cross-reactive and sustained immune response after SARS-CoV-2 infection^41^, the observed pulmonary dysfunction in our study is an unexpected finding.

This trial has several limitations. We did not compare our measurements to another reference standard, such as ventilation-perfusion scintigraphy, spirometry or body plethysmography. However, most of these modalities either use ionization radiation, are invasive or require active cooperation. In addition, validation of our imaging approach has been described previously^26,29^. The functional LF-MRI in our study used free-breathing all intervals, which was feasible in 93% of the pediatric post-acute Covid-19 patients starting from 5 years of age. Given our technical setup, measurements of functional lung parenchyma had systematically lower values as reported before,^28,30,42^ which does not alter our comparisons between the investigated groups. Further limitations include the lack of longitudinal data as well as the lower number of healthy controls. Finally, a selection bias could exist, as a majority of families with acute or post-acute symptomatic children and higher disease burden might have participated in the study. However, we obtained a balanced cohort with 24 (44%) long Covid patients, putting this potential bias into perspective.

In summary, we report persisting pulmonary dysfunction both in pediatric patients recovered from Covid-19 and long Covid patients. The further course and outcome of the observed changes currently remains unclear. Our results warrant further surveillance of persistent pulmonary damage in pediatrics and adolescents after SARS-CoV-2 infection. Given the translatability of the technology, these functional imaging approaches can be rapidly adopted to clinical routine care.

## Data Availability

All data produced in the present study are available upon reasonable request to the authors.

## Acknowledgments

We thank the Imaging Science Institute Erlangen for providing us with measurement time and technical support. Many thanks to Natalie Wendisch, Kristin Prypilla und Annika Maischberger for technical assistance during MR imaging and the staff of the social pediatrics center at the University Hospital Erlangen during patient recruitment.

The present work was performed in (partial) fulfillment of the requirements for obtaining the degree „Dr. med.” for L.T. and in (partial) fulfillment of the requirements for obtaining the degree „Dr. rer. biol. hum.” for A.P.R.

## Data availability statement

The data sets generated during and/or analyzed during the current study are available from the corresponding author on reasonable request as follows:

Individual participant data will not be available. Study Protocol and Statistical Analysis Plan will be available. All data will be available beginning 9 months and ending 36 months following article publication. The data will be available to researchers who provide a methodologically sound proposal. The data will be available for individual participant data meta-analysis, only. Proposals may be submitted up to 36 months following article publication. After 36 months the data will be available in our university’s data warehouse but without investigator support other than deposited metadata. Information regarding submitting proposals and accessing data may be found at https://www.uk-erlangen.de. Restrictions may apply due to patient privacy and the General Data Protection Regulation.

## Code availability statement

No previously unreported custom computer code or algorithm was used to establish results in this paper.

