## Supplementary Appendix for "Persisting pulmonary dysfunction in pediatric post-acute Covid-19"

This appendix has been provided by the authors to give readers additional information about the work.

### Supplementary methods

**Roche Elecsys® Anti-SARS-CoV-2 assay and Anti-SARS-CoV-2 -S assay**

All blood samples were centrifuged and stored as serum at -15 degrees.. . For qualitative testing we used the electrochemiluminescence immunoassay (ECLIA) Elecsys® Anti-SARS-Cov-2 assay according to the manufactures protocol. Briefly, it uses a recombinant protein representing the nucleocapsid (N) antigen in a double-antigen sandwich assay format, detecting antibody titer of all antibody isotypes (IgM, IgA, IgG). Test results exceeding 1.0 were interpreted as reactive, following the manufacturer’s suggested Cut-off-index. Assay sensitivity was previously evaluated to have a sensitivity of 99.5% 14 days after the first positive PCR results (95% CI: 97,0%-100%) and a specificity of 99.8% (95% CI: 99.69%-99.88%)”.^1^ All samples were further assessed on the same cobas e 601 module with the Elecsys® Anti-SARS-CoV-2-S immunoassay (ACOV2S). Anti-SARS-CoV-2 S is an immunoassay for the in vitro quantitative determination of antibodies to the SARS-CoV-2 spike (S) protein in human serum and plasma. It uses a recombinant protein representing the receptor binding domain of the spike protein enabling the identification of highly affine SARS-CoV-2 IgM and IgG antibodies using a double antigen sandwich immunoassay. All samples were processed according to the manufacturer’s instructions. Test results exceeding 0.8U/ml were considered reactive, following the manufacturer’s suggested cut-off-index. Values between 0.40–250 U/mL represent the linear range. Samples above 250 U/mL were automatically diluted into the linear range of the assay (realized dilutions in this study: 1:10 or 1:100) with Diluent Universal (Roche Diagnostics, Mannheim, Germany). The analyzer automatically multiplies diluted results with the dilution factor, which in the applied setting enabled an upper limit of quantification of 25000 U/mL for these analyses. The assay demonstrated a sensitivity of 98.9% (95% CI: 98.1%-99.3%) 14 days after the first positive PCR result and a specificity of 99.96% (95% CI: 99.91%-100%).^2^

The assigned U/mL are equivalent to Binding Antibody Units (BAU)/mL as defined by the first World Health Organization (WHO) International Standard for anti-SARS-CoV-2 immunoglobulin (NIBSC code 20/136). The reported results in U/mL can be directly compared to other studies or results in BAU/mL. No conversion of units is required.

Serum Interleukin 6 (IL-6) was measured by electrochemiluminescence immunoassay (Elecsys IL-6, Cobas e601). C-reactive protein (CrP) was measured using a standard immunoturbidometric assay on the Cobas c501 system (Roche Diagnostics).

Complete blood counts were performed using Sysmex XE-2100 (Sysmex, Kobe, Japan) analyzers.

### Supplementary Tables

**Table S1** – Functional low-field magnetic resonance imaging parameter

| **Parameter** | **Explanation** |
| --- | --- |
| Mean FVL Correlation | Ventilation derived by cross-correlation |
| VDP_Total_ | Ventilation defected percentage |
| QDP_Total_ | Perfusion defected percentage |
| VQM_Defect_ | Ventilation/perfusion match (defect) |
| VQM_Non-defect_ | Ventilation/perfusion match |
| VDP_Exclusive_ | Exclusive ventilation defect percentage, no concurrent Q defect, based on regional ventilation |
| VDP_FVL,Exclusive_ | Exclusive ventilation defect percentage, no concurrent Q defect, based on FVL correlation metric |
| VDP_FVL_ | Ventilation defect percentage, based on flow-volume loop correlation metric |
| QDP_Exclusive_ | Exclusive perfusion defect percentage, no concurrent V defect, based on normalized perfusion |
| VQM_Defect, FVL_ | VQ Match (defect) based on Q and FVL |
| VQM_Non-defect, FVL_ | VQ Match (non-defect) based on Q and FVL |

**Table S2** – Acute and post-acute symptoms of the study population

| **Characteristic** | **Healthy control**  **(N=9)** | **Post Covid-19**  **(N=54)** |
| --- | --- | --- |
| **Acute Symptoms during SARS-CoV-2 infection – no. (%)** |  |  |
| Headache | N/A | 29 (54) |
| Rhinitis | N/A | 29 (54) |
| Sore throat | N/A | 23 (43) |
| Cough | N/A | 25 (46) |
| Shortness of breath | N/A | 8 (15) |
| Pneumonia | N/A | 0 (0) |
| Fever | N/A | 22 (41) |
| Anosmia | N/A | 13 (24) |
| Ageusia | N/A | 17 (31) |
| Fatigue | N/A | 10 (19) |
| Diarrhea | N/A | 1 (2) |
| Limb pain | N/A | 5 (9) |
| **Post-acute Symptoms after SARS-CoV-2 infection – no. (%)** |  |  |
| Headache | N/A | 5 (9) |
| Dyspnea | N/A | 15 (28) |
| Pneumonia | N/A | 1 (2) |
| Fever | N/A | 0 (0) |
| Anosmia | N/A | 4 (7) |
| Ageusia | N/A | 1 (2) |
| Fatigue | N/A | 4 (7) |
| Impaired attention | N/A | 6 (11) |
| Limb pain | N/A | 1 (2) |
| Shortness of breath | N/A | 16 (30) |

**Table S3**– Quantitative measurements of free breathing phase-resolved function lung magnetic resonance imaging*

| **Parameter** | **Healthy control**  **(N=9)** | **Recovered**  **(N=30)** | **P Value†** | **Long Covid**  **(N=24)** | **P Value†** |
| --- | --- | --- | --- | --- | --- |
| Mean FVL Correlation | 0.94±0.03 | 0.89±0.06 | 0.17 | 0.88±0.10 | 0.25 |
| VDP_Exclusive_ - % | 12.3±3.4 | 18.4±6.8 | 0.04 | 19.1±8.2 | 0.03 |
| VDP_FVL,Exclusive_ - % | 9.2±3.9 | 14.0±7.5 | 0.26 | 14.9±9.4 | 0.24 |
| VDP_FVL_ - % | 9.8±4.1 | 17.4±9.5 | 0.12 | 20.1±13.3 | 0.06 |
| QDP_Exclusive_ - % | 6.0±4.3 | 14.9±14.9 | 0.58 | 17.1±13.6 | 0.15 |
| VQM_Defect, FVL_ - % | 0.6±0.8 | 3.4±4.2 | 0.29 | 5.1±7.9 | 0.04 |
| VQM_Non-defect, FVL_ - % | 84.7±7.4 | 67.4±20.9 | 0.06 | 62.2±22.2 | 0.02 |

* Plus–minus values are means ±SD.

† P values tested with Kruskal-Wallis test and Dunn’s post test for multiple comparisons and given in comparison to healthy controls.

**Table S4** – Diagnostic characteristics of functional low-field magnetic resonance imaging parameters*

| **Parameter** | **AUC** | **95%CI** | **P value** | **Likelihood** | **Cut-off** | **Sensitivity** | **Specificity** | **No. of patients detected** |
| --- | --- | --- | --- | --- | --- | --- | --- | --- |
| **Ventilation defected percentage (VDP_total_)** |  | | | | | |  |  |
| Recovered | 0.83 | 0.70-0.96 | 0.003 | 6.5 | >17.5% | 72.4% | 88.9% | 21/29 |
| Long Covid | 0.88 | 0.76-0.99 | <0.001 | 6.8 | >17.5% | 76.0% | 88.9% | 19/25 |
| Combined | 0.85 | 0.75-0.85 | <0.001 | 6.7 | >17.5% | 74.1% | 88.9% | 40/54 |
| **Perfusion defected percentage (QDP_Total_)** |  | | | | | |  |  |
| Recovered | 0.67 | 0.50-0.84 | 0.13 | 5.0 | >9.9% | 55.2% | 88.9% | 16/29 |
| Long Covid | 0.74 | 0.58-0.91 | 0.03 | 6.1 | >9.5% | 68.0% | 88.9% | 17/25 |
| Combined | 0.70 | 0.57-0.84 | 0.05 | 5.5 | >9.5% | 61.1% | 88.9% | 33/54 |
| **Ventilation/perfusion mismatch (VQM_Defect_)** |  | | | | | |  |  |
| Recovered | 0.77 | 0.62-0.93 | 0.01 | 5.6 | >0.85% | 62.1% | 88.9% | 18/29 |
| Long Covid | 0.88 | 0.75-1.00 | <0.001 | 6.8 | >0.85% | 76.0% | 88.9% | 19/25 |
| Combined | 0.82 | 0.69-0.95 | 0.002 | 6.2 | >0.85% | 68.5 | 88.9% | 37/54 |
| **Ventilation/perfusion match (VQM_Non-defect_)** |  | | | | | |  |  |
| Recovered | 0.84 | 0.70-0.98 | 0.002 | 7.1 | <77.4% | 79.3% | 88.9% | 26/29 |
| Long Covid | 0.88 | 0.75-1.00 | <0.001 | 7.2 | <76.4% | 80.0% | 88.9% | 21/25 |
| Combined | 0.86 | 0.74-0.97 | <0.001 | 7.2 | <77.4% | 79.6% | 88.9% | 47/54 |

* Plus–minus values are means ±SD.

### Supplementary Figures

**Figure S1** – Morphologic low-field magnetic resonance imaging of pulmonary consolidations in one patient.

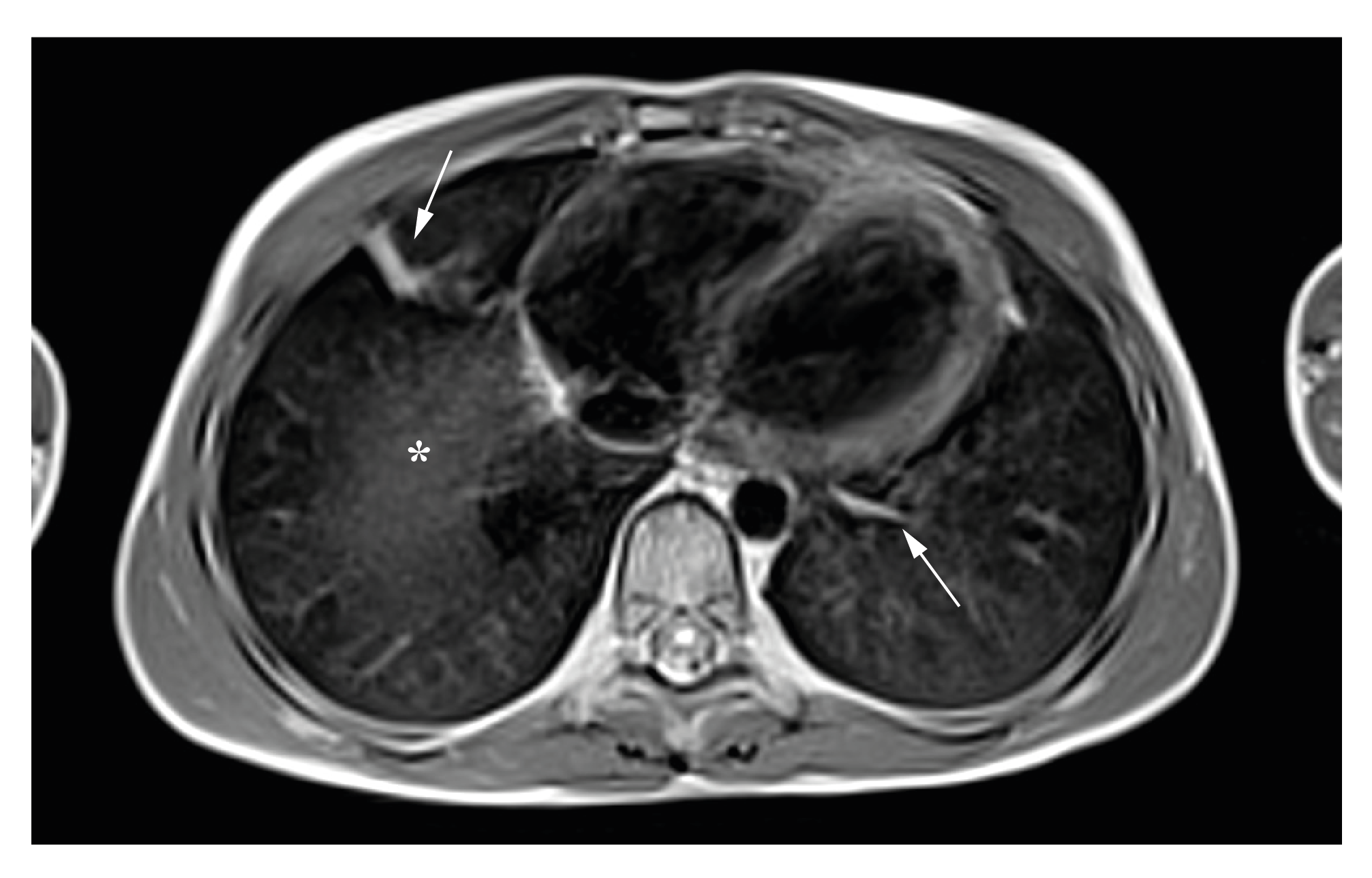

Arrows indicate pulmonary consolidations. Asterics marks a partial volume effect caused by adjacent liver parenchyma.

**Figure S2** – Low-field magnet resonance imaging measurements in post-acute Covid-19 patients (n=54) vs. healthy controls (n=9)

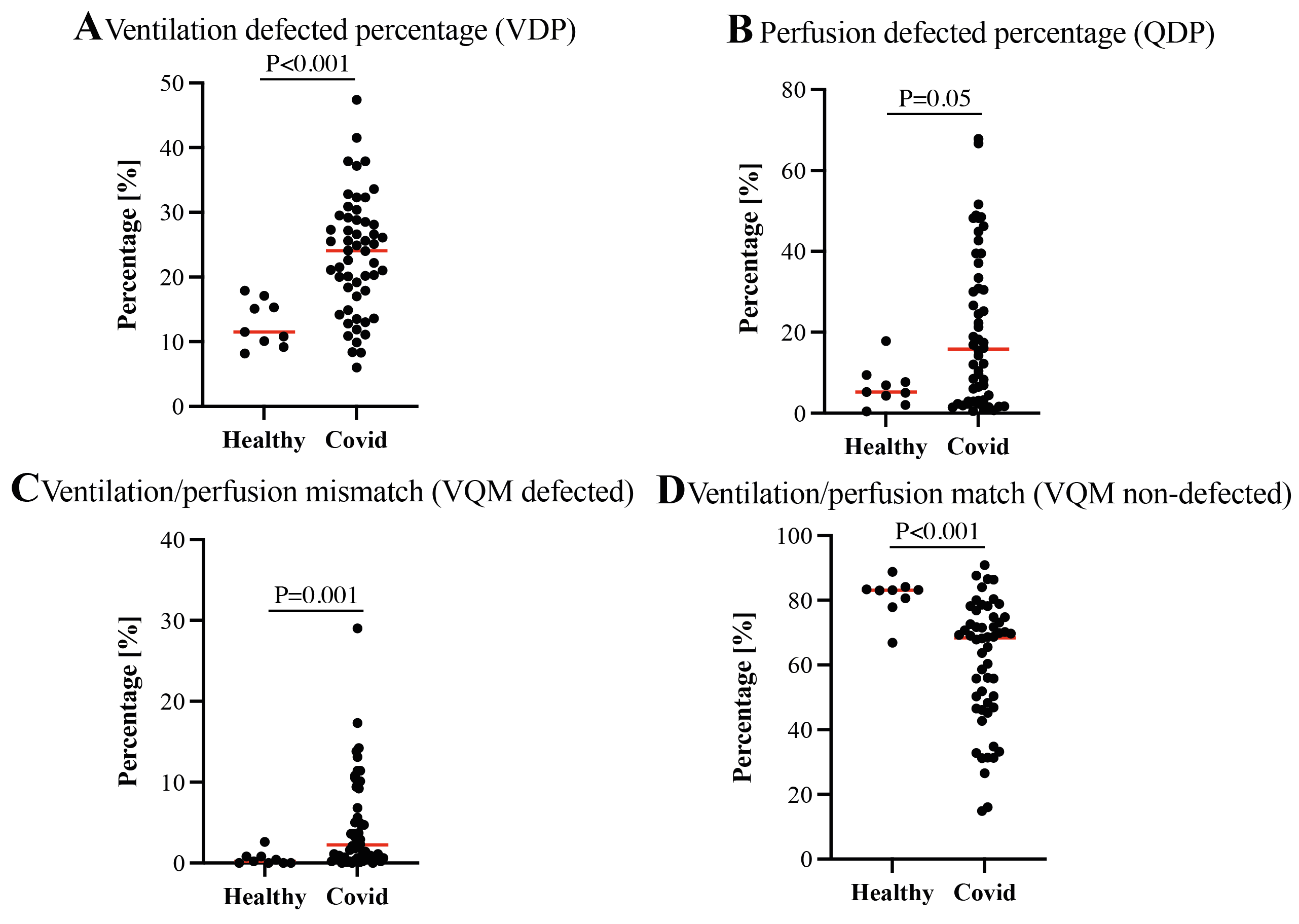

All test performed with non-parametric Mann-Whitney test.

**Figure S3** – Morphologic low-field magnetic resonance imaging of healthy control, recovered and long Covid patient

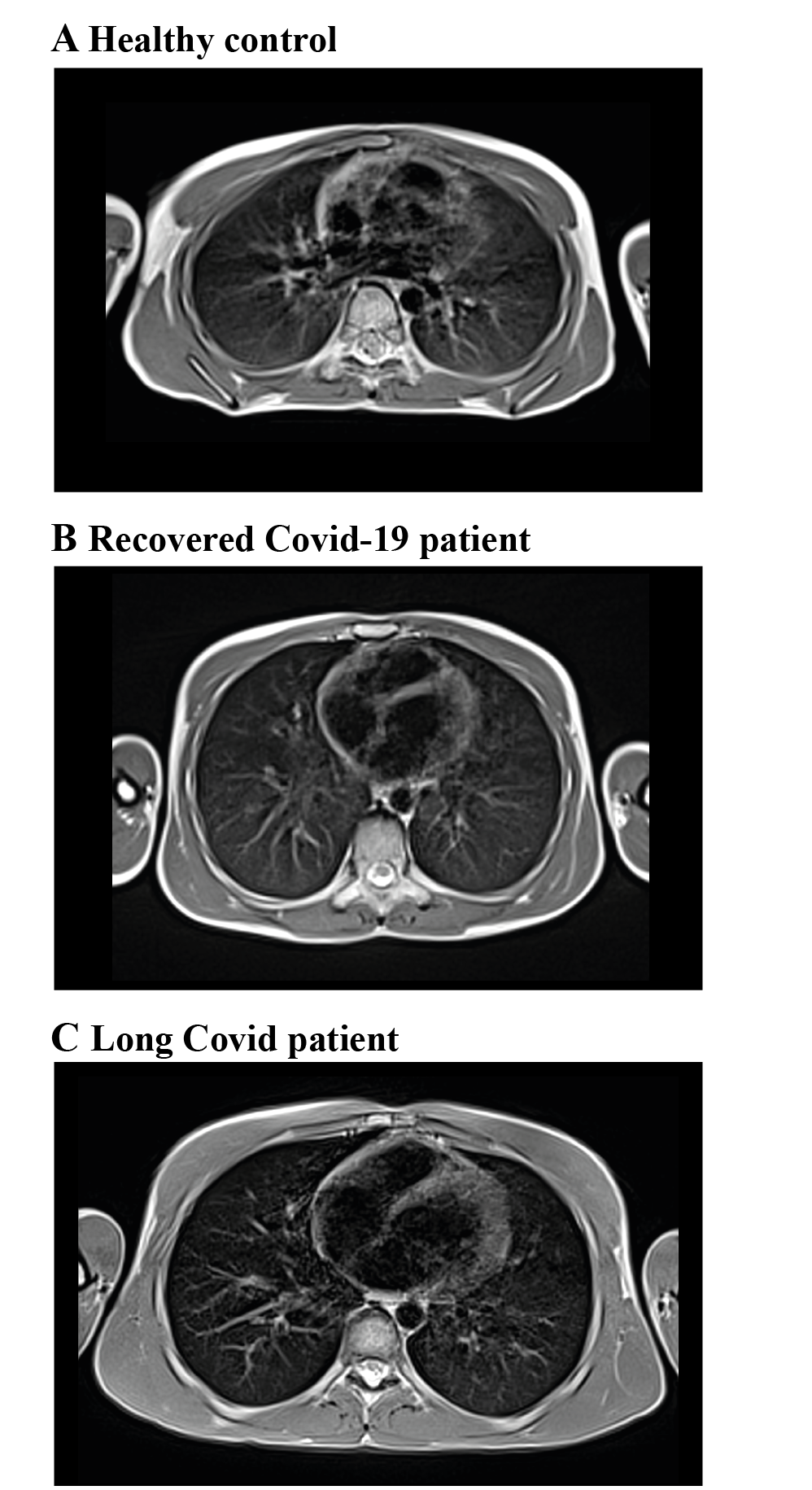

**Figure S4** – Low-field magnet resonance imaging findings in healthy controls

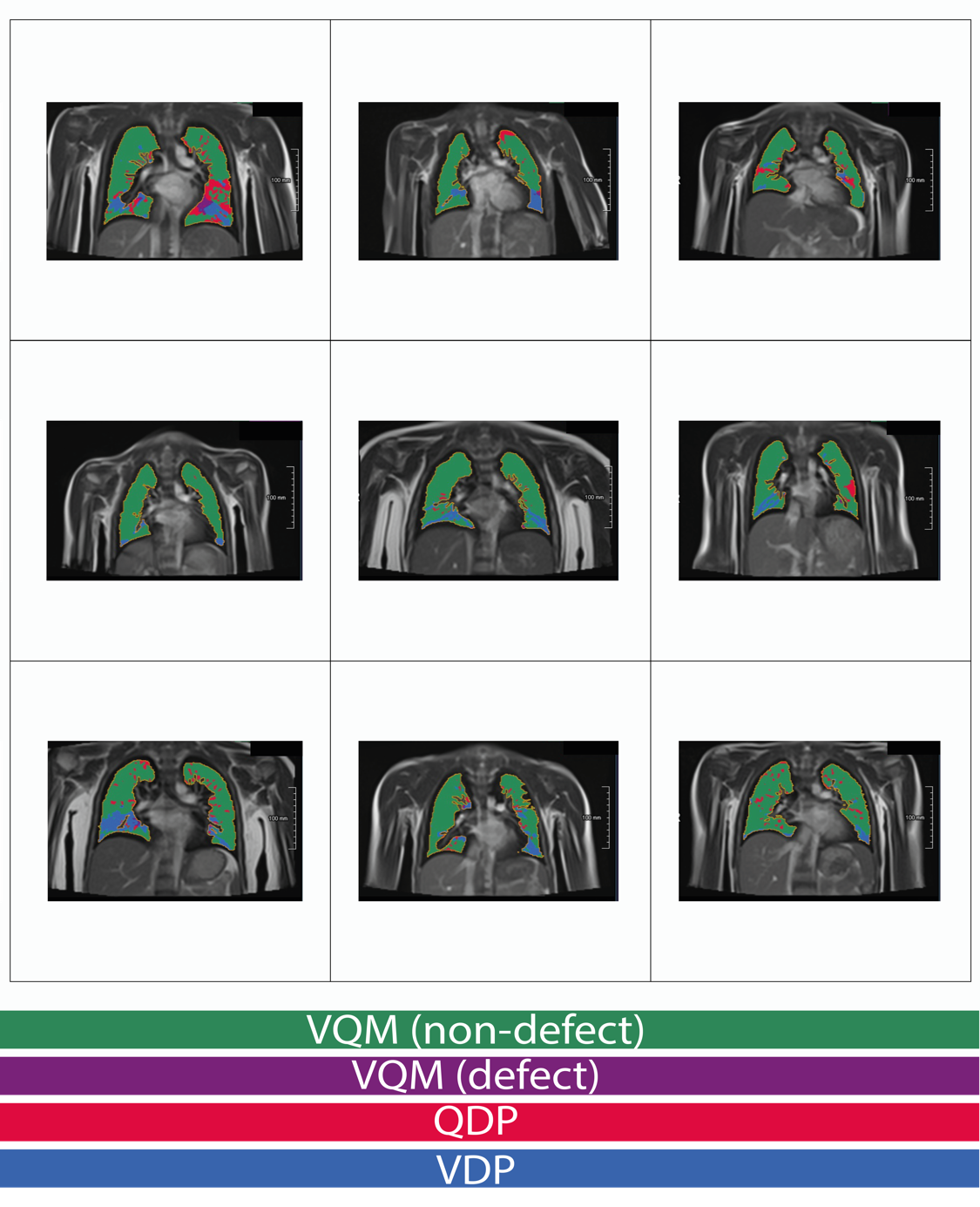

VQM (non-defected) = ventilation/perfusion match; VQM (defect) = ventilation/perfusion mismatch; QDP (defect) = perfusion defected area; VDP (defect) = ventilation defected area

**Figure S5** – Low-field magnet resonance imaging findings in recovered Covid-19 patients

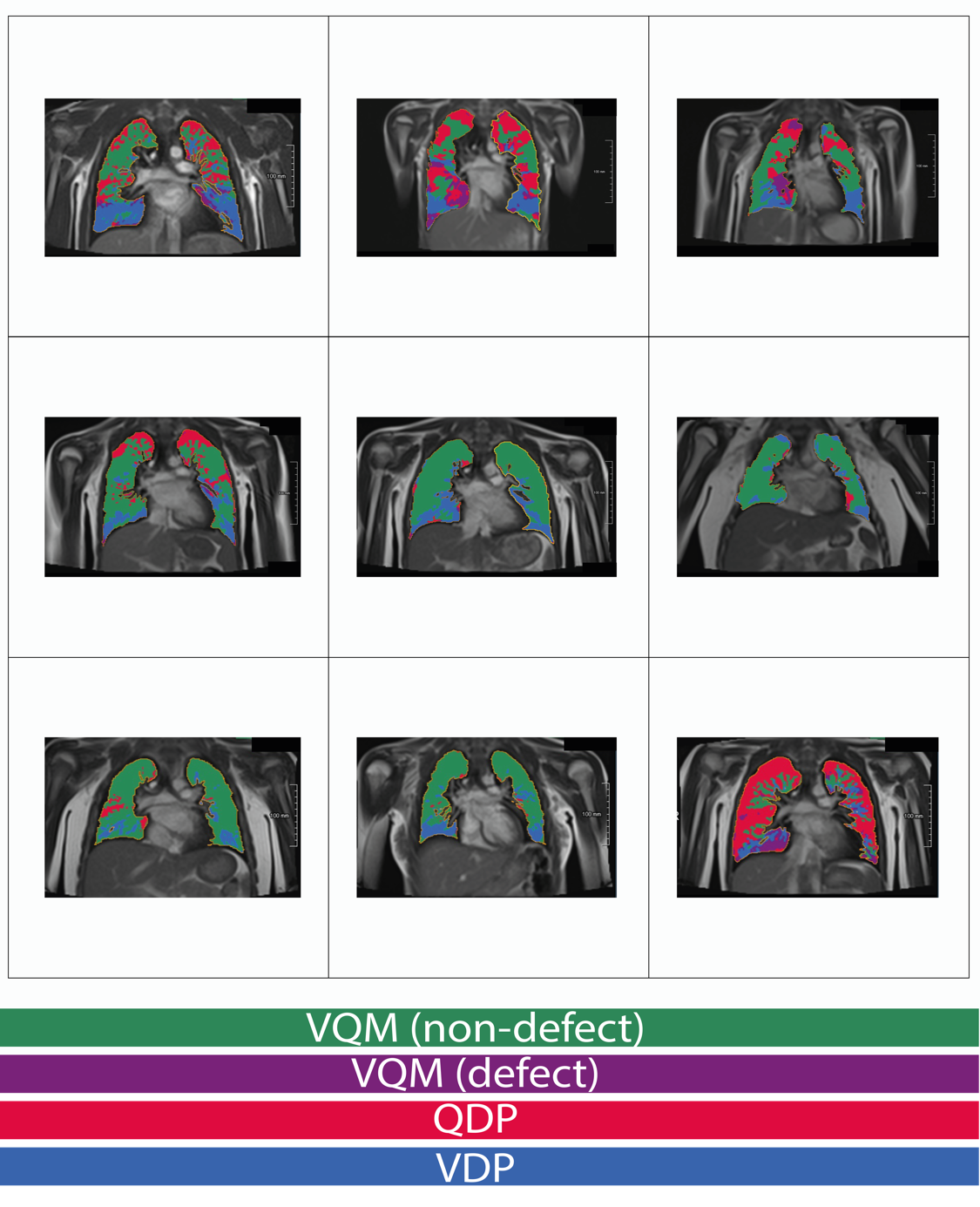

VQM (non-defected) = ventilation/perfusion match; VQM (defect) = ventilation/perfusion mismatch; QDP (defect) = perfusion defected area; VDP (defect) = ventilation defected area

**Figure S6** – Low-field magnet resonance imaging findings in recovered Covid-19 patients

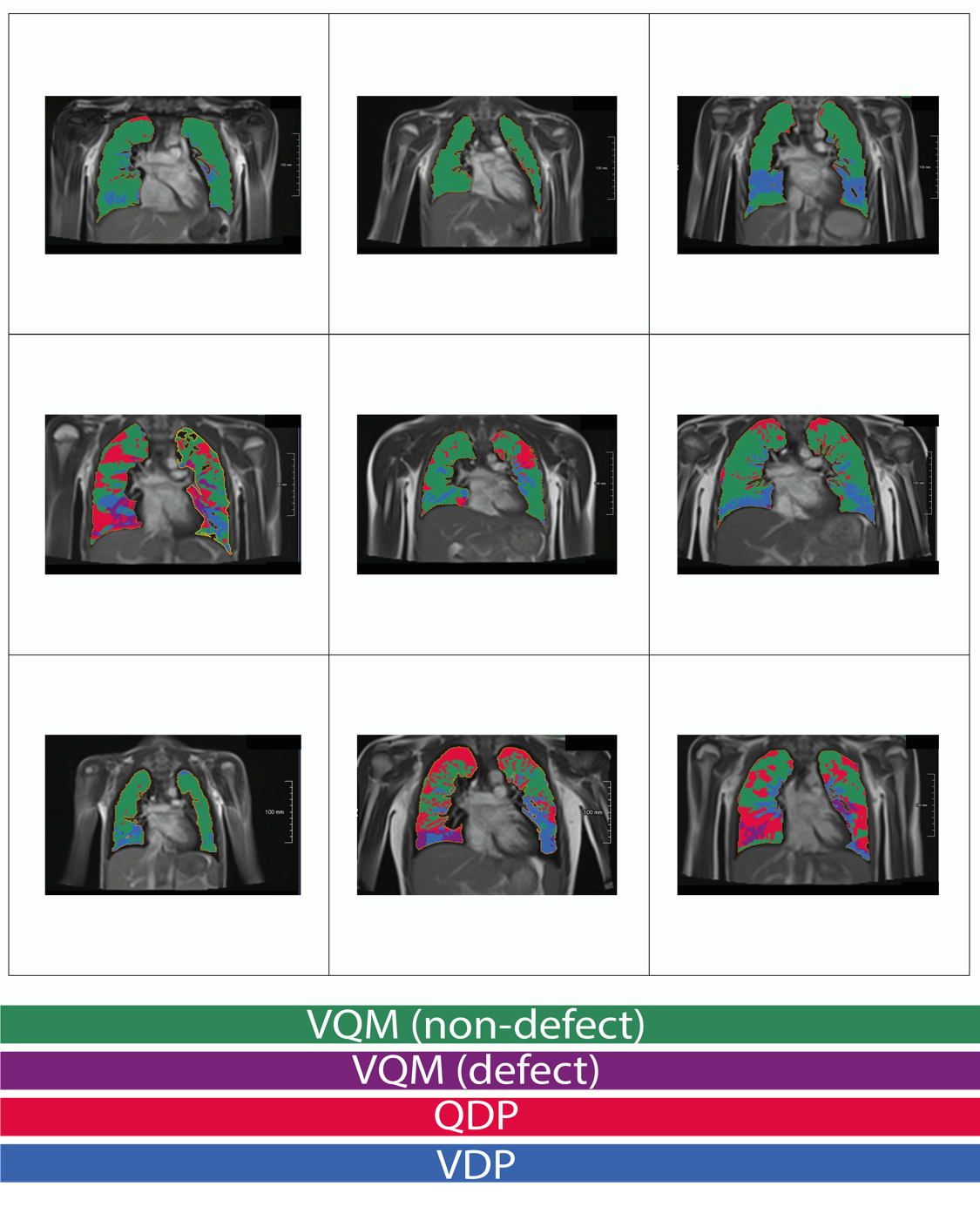

VQM (non-defected) = ventilation/perfusion match; VQM (defect) = ventilation/perfusion mismatch; QDP (defect) = perfusion defected area; VDP (defect) = ventilation defected area

**Figure S7** – Low-field magnet resonance imaging findings in recovered Covid-19 patients

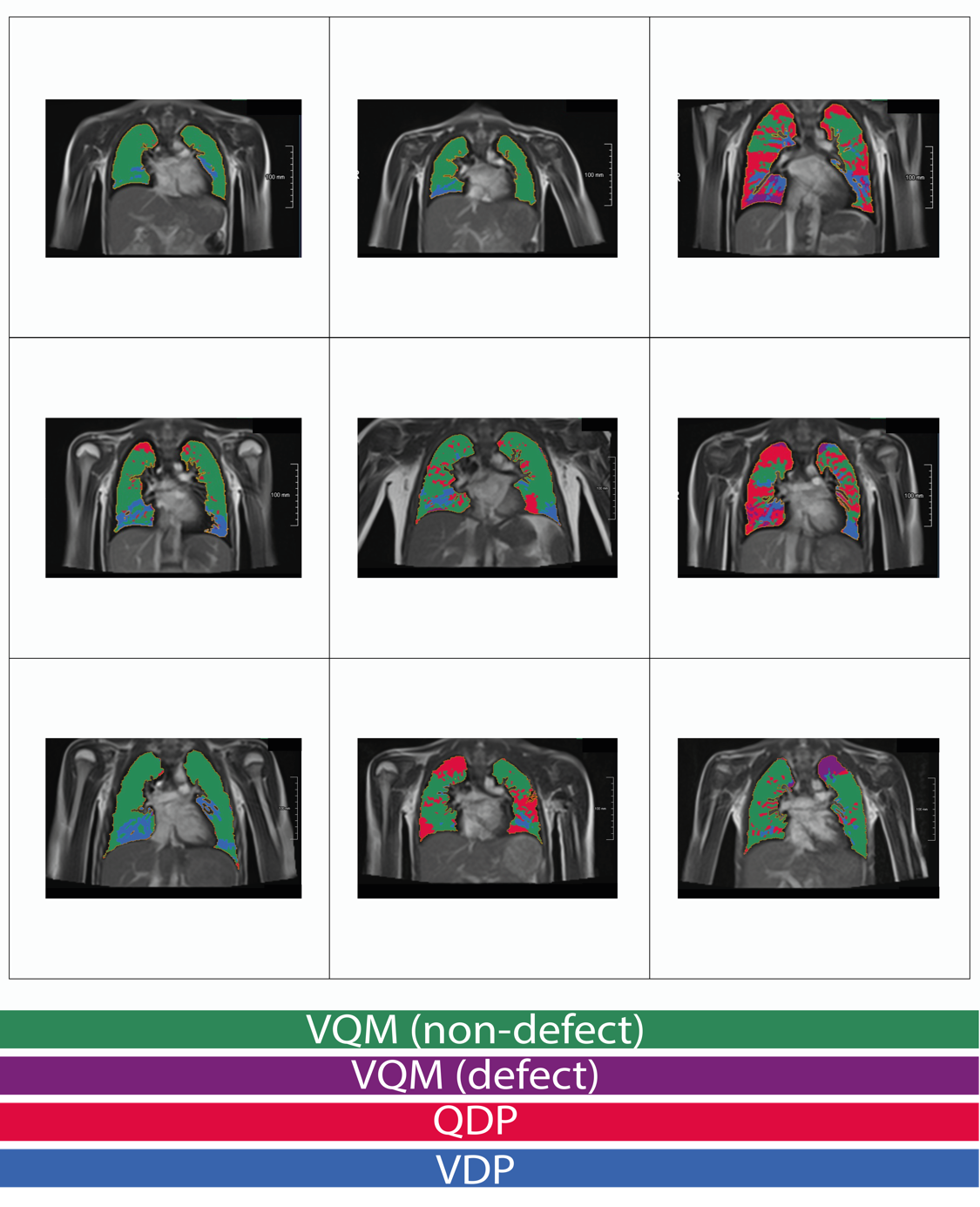

VQM (non-defected) = ventilation/perfusion match; VQM (defect) = ventilation/perfusion mismatch; QDP (defect) = perfusion defected area; VDP (defect) = ventilation defected area

**Figure S8** – Low-field magnet resonance imaging findings in recovered Covid-19 patients

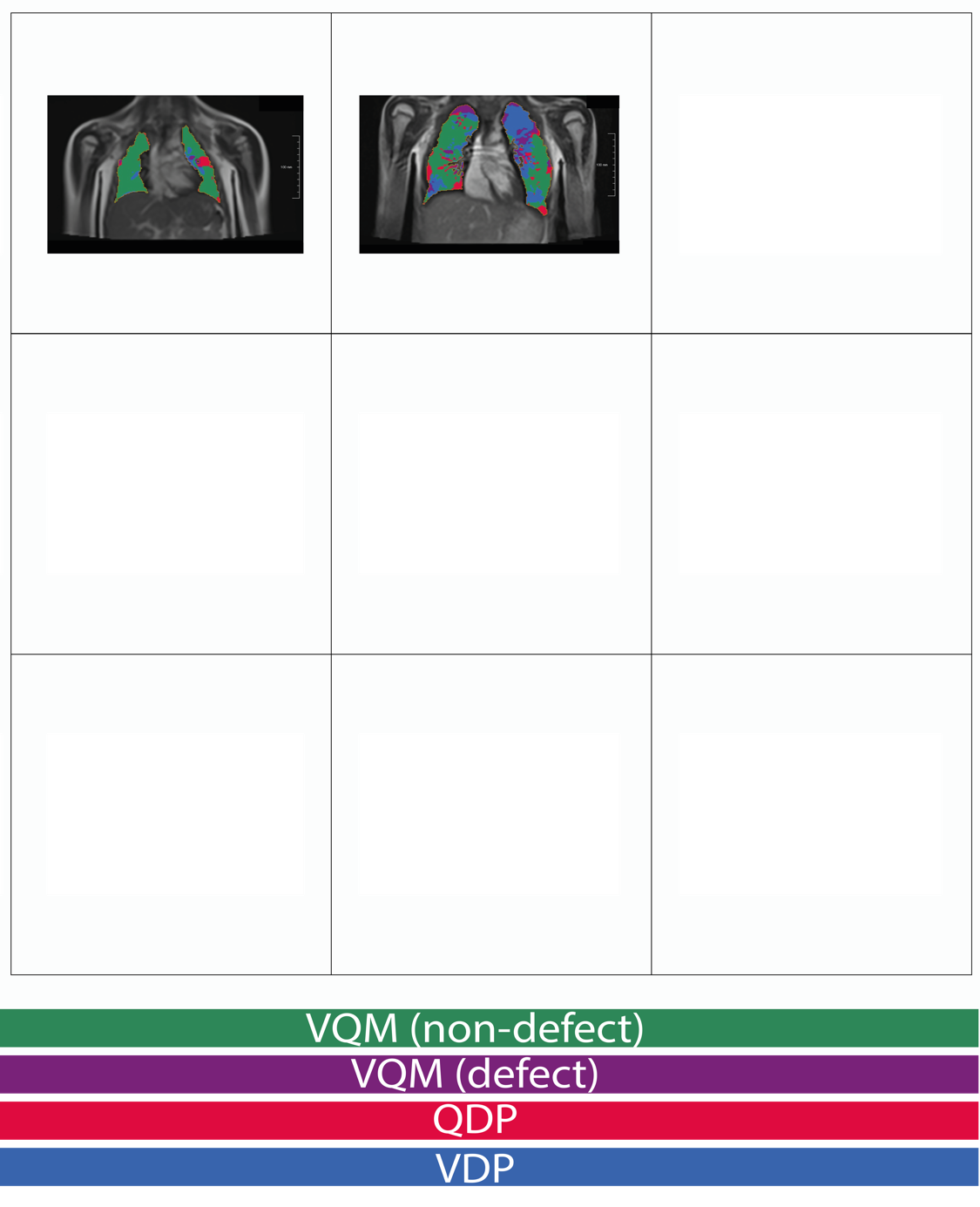

VQM (non-defected) = ventilation/perfusion match; VQM (defect) = ventilation/perfusion mismatch; QDP (defect) = perfusion defected area; VDP (defect) = ventilation defected area

**Figure S9** – Low-field magnet resonance imaging findings in long Covid-19 patients

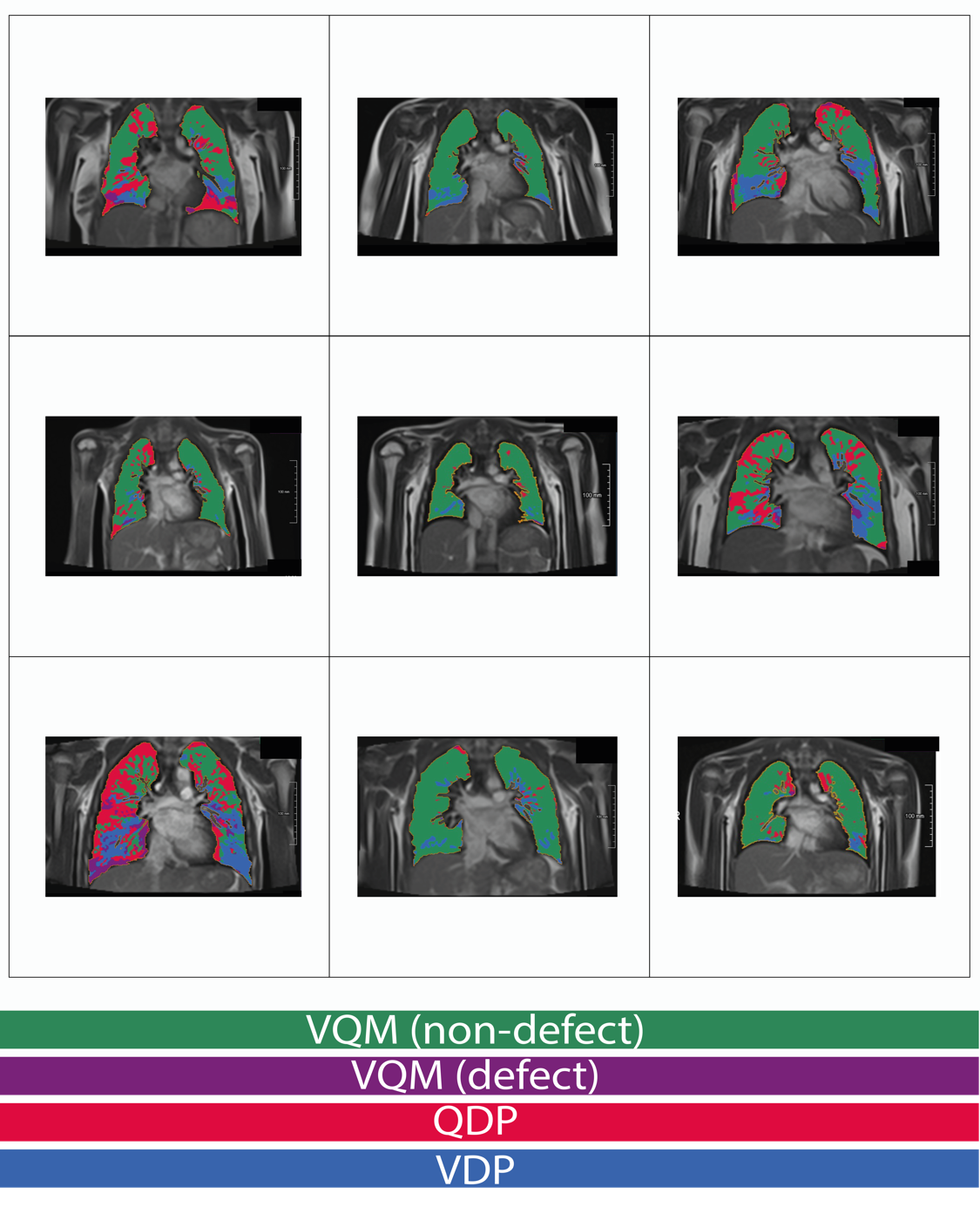

VQM (non-defected) = ventilation/perfusion match; VQM (defect) = ventilation/perfusion mismatch; QDP (defect) = perfusion defected area; VDP (defect) = ventilation defected area

**Figure S10** – Low-field magnet resonance imaging findings in long Covid-19 patients

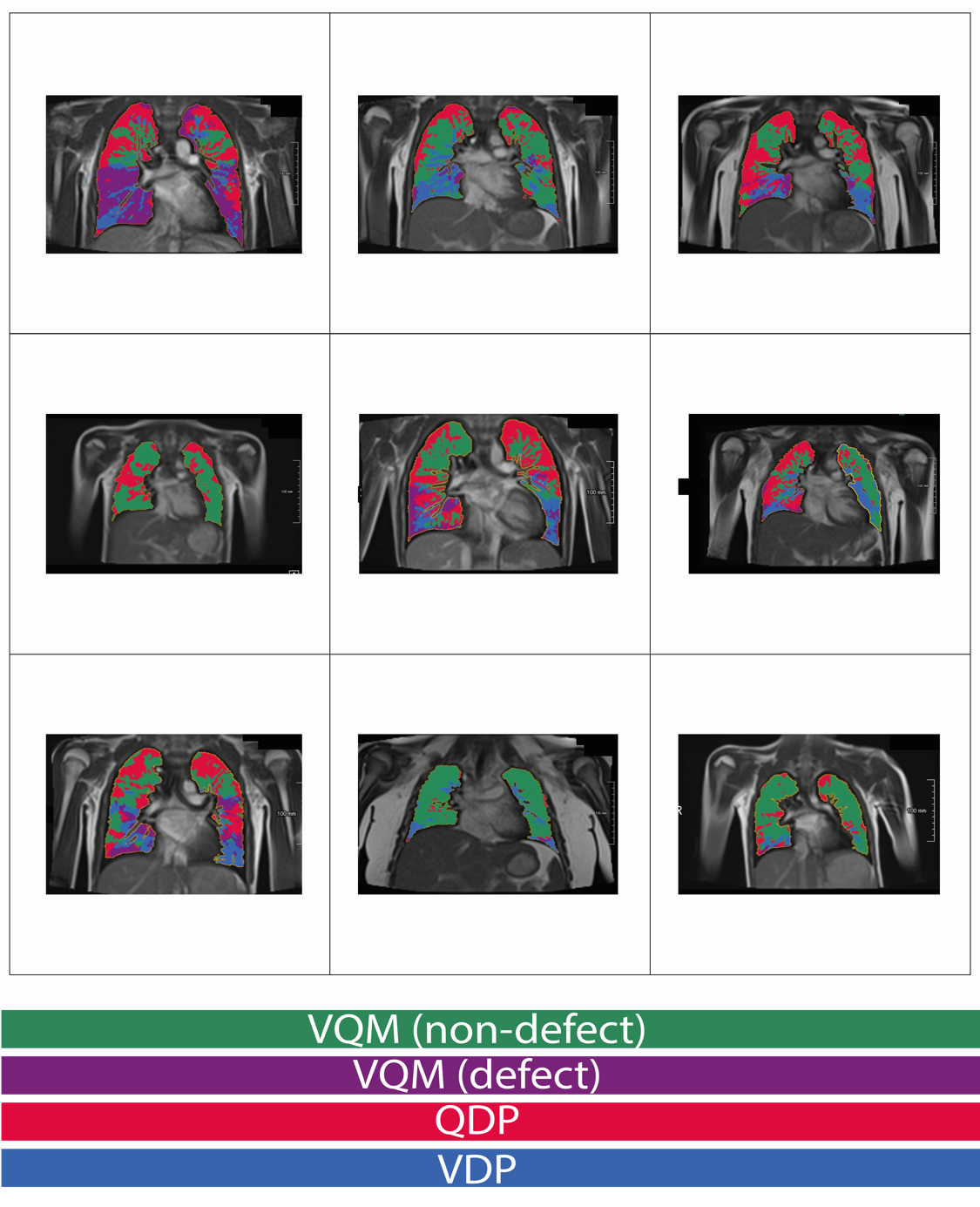

VQM (non-defected) = ventilation/perfusion match; VQM (defect) = ventilation/perfusion mismatch; QDP (defect) = perfusion defected area; VDP (defect) = ventilation defected area

**Figure S11** – Low-field magnet resonance imaging findings in long Covid-19 patients

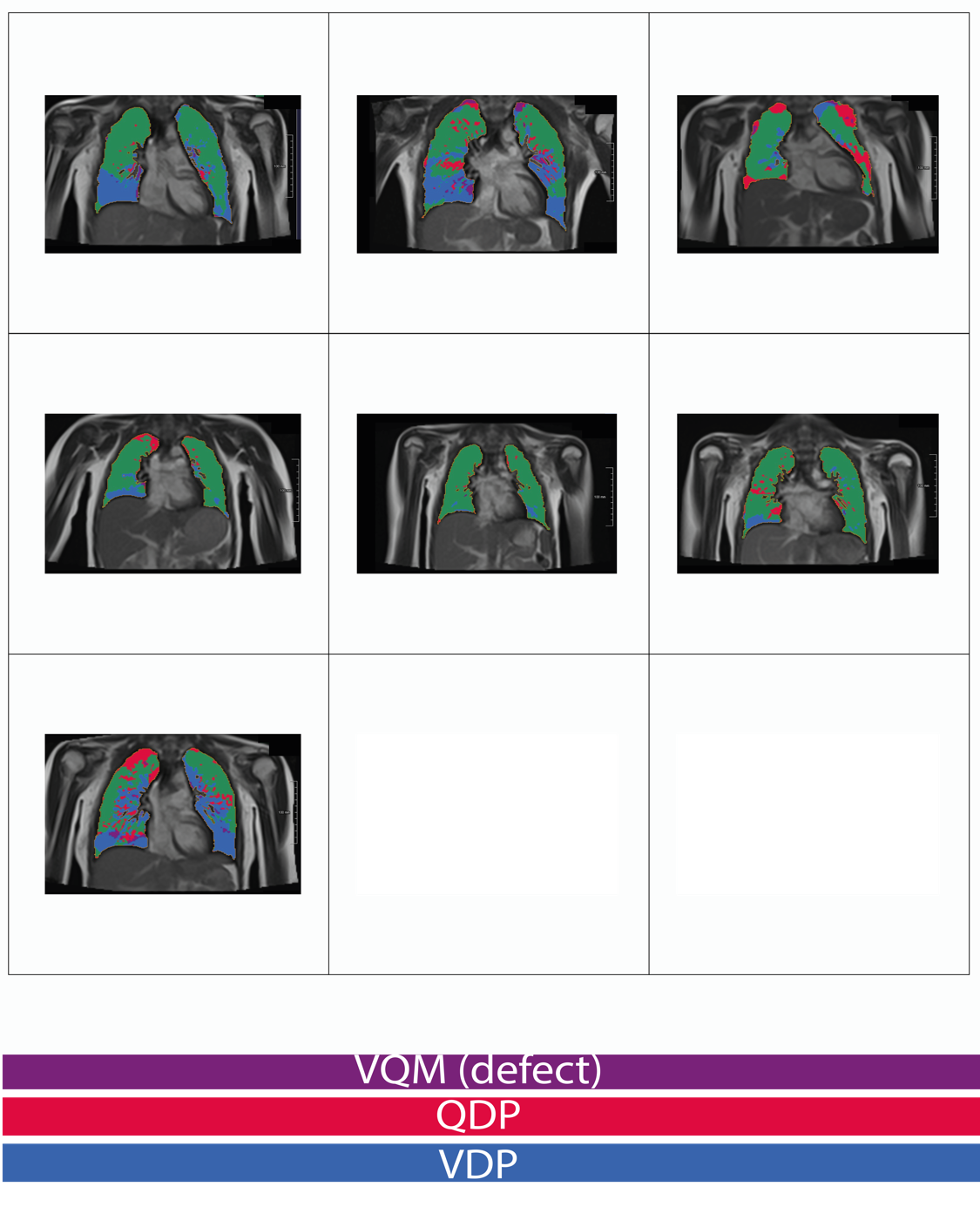

VQM (non-defected) = ventilation/perfusion match; VQM (defect) = ventilation/perfusion mismatch; QDP (defect) = perfusion defected area; VDP (defect) = ventilation defected area

**Figure S12** – Low-field magnet resonance imaging measurements in healthy controls (n=9), recovered (n=29) and long Covid (n=24) patients.

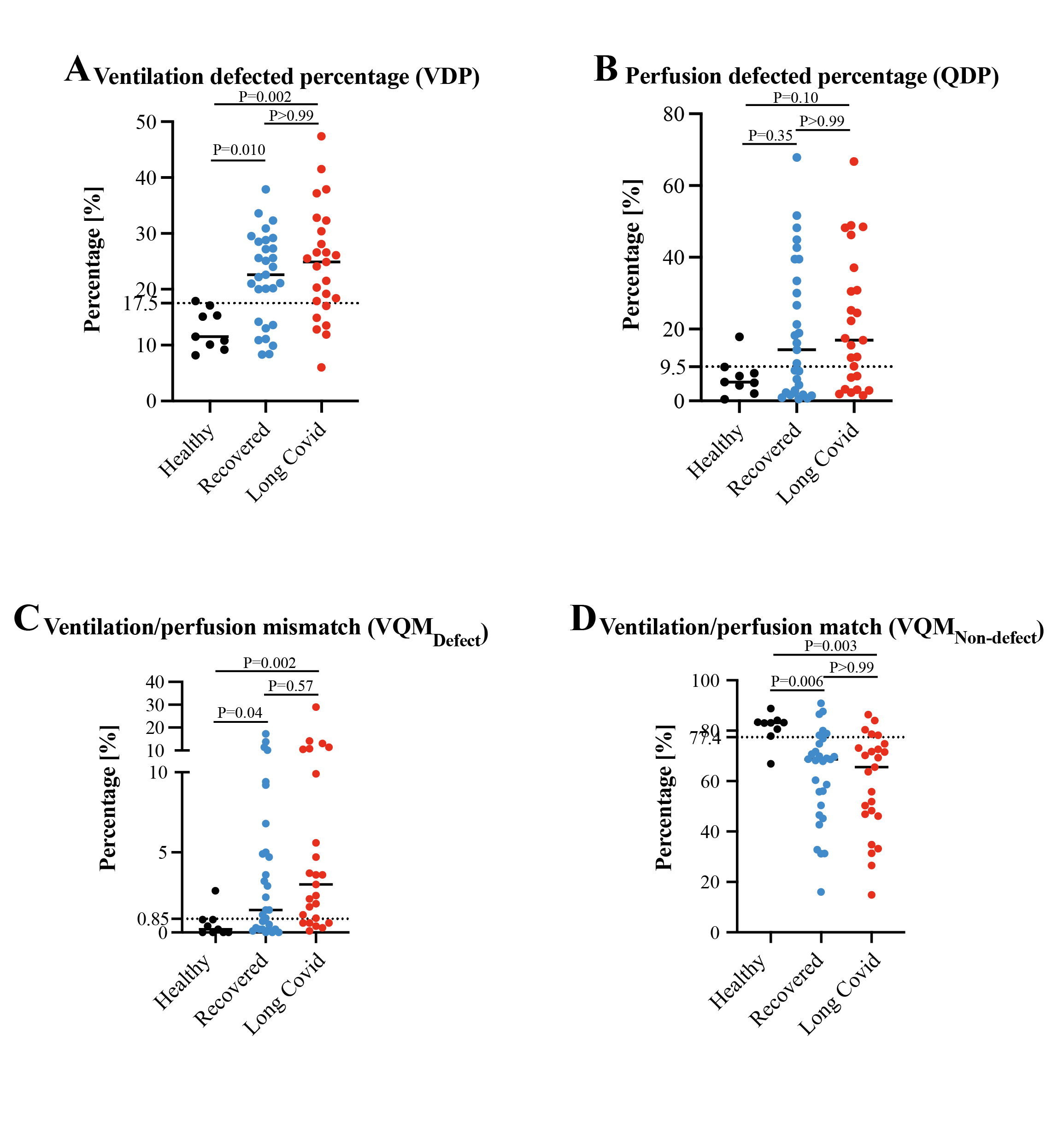

Dashed lines indicate cut-off derived from ROC analyses. P values tested with Kruskal-Wallis test and Dunn’s post test for multiple comparisons and given in comparison to healthy controls.

2.	Fact sheet, ElecsysT Anti-SARS-CoV-2 S. (<https://assets.cwp.roche.com/f/94122/x/379ebe6732/factsheet-elecsys-anti-sars-cov-2-s_v1.pdf>).
